# Severity, criticality, and fatality of the SARS-CoV-2 Beta variant

**DOI:** 10.1101/2021.08.02.21261465

**Authors:** Laith J. Abu-Raddad, Hiam Chemaitelly, Houssein H. Ayoub, Hadi M. Yassine, Fatiha M. Benslimane, Hebah A. Al Khatib, Patrick Tang, Mohammad R. Hasan, Peter Coyle, Sawsan AlMukdad, Zaina Al Kanaani, Einas Al Kuwari, Andrew Jeremijenko, Anvar Hassan Kaleeckal, Ali Nizar Latif, Riyazuddin Mohammad Shaik, Hanan F. Abdul Rahim, Gheyath K. Nasrallah, Mohamed Ghaith Al Kuwari, Adeel A. Butt, Hamad Eid Al Romaihi, Mohamed H. Al-Thani, Abdullatif Al Khal, Roberto Bertollini

## Abstract

Severity (acute-care hospitalization), criticality (ICU hospitalization), and fatality of SARS-CoV-2 Beta (B.1.351) variant was investigated through case-control studies applied to complete national cohorts of infection, disease, and death cases in Qatar. Compared to Alpha (B.1.1.7) variant, odds of progressing to severe disease were 1.24-fold (95% CI: 1.11-1.39) higher for Beta. Odds of progressing to critical disease were 1.49-fold (95% CI: 1.13-1.97) higher. Odds of COVID-19 death were 1.57-fold (95% CI: 1.03-2.43) higher. Findings highlight risks to healthcare systems, particularly to intensive care facilities and resources, with increased circulation of Beta.

## Introduction

Commencing in mid-January, 2021, Qatar experienced a severe acute respiratory syndrome coronavirus 2 (SARS-CoV-2) Alpha (B.1.1.7) variant wave that peaked in the first week of March [1–5], but was immediately followed by a Beta (B.1.351) variant wave that peaked in the first week of April [1–5]. This created a unique epidemiologic situation that allowed comparative assessment of the severity, criticality, and fatality of these two variants.

## Methods

We investigated severity (acute-care hospitalization) [6], criticality (ICU hospitalization) [6], and fatality [7] of both variants through eight case-control studies applied to the complete national cohorts of SARS-CoV-2 infections, Coronavirus Disease 2019 (COVID-19) disease cases, and COVID-19 deaths in Qatar, a country with diverse demographics where 89% of the population comprises expatriates from over 150 countries [8]. Data on polymerase chain reaction (PCR) testing and clinical characteristics were extracted from the national, federated COVID-19 databases that have captured all SARS-CoV-2–related data since the start of the epidemic. These databases were retrieved from the integrated nationwide digital-health information platform (universal healthcare system), and include all records of PCR testing, antibody testing, vaccinations, COVID-19 hospitalizations, infection severity classification, and COVID-19 deaths. Databases are complete at the national level with no missing information.

Records of PCR testing and clinical data for hospitalized COVID-19 patients were examined. Details of the laboratory methods for PCR testing are found in Supplementary Text 1. Each person that had a positive PCR test result and hospital admission was subject to an infection severity assessment every three days until discharge or death. Individuals who progressed to COVID-19 disease between the time of the positive PCR test result and the end of the study were classified based on their worst outcome, starting with death [7], followed by critical disease [6], and then severe disease [6].

Cases in the case-control studies were persons who progressed to COVID-19 severe disease, critical disease, or death. Controls were persons with asymptomatic or mild SARS-CoV-2 infections. Cases and controls were matched at a ratio of 1:3 by 10-year age group, sex, and bi-weekly interval of the PCR diagnosis date. Every case in Qatar that met the inclusion criteria and that could be matched to a control was included in the study. Classification of case severity, criticality, and fatality followed the World Health Organization guidelines [6, 7], and assessments were made by trained medical personnel through individual chart reviews. Details of the COVID-19 severity, criticality, and fatality classification are found in Supplementary Text 2.

From January 18 until February 15, 2021, the Alpha variant wave expanded rapidly and weekly rounds of viral genome sequencing [1–4] of randomly collected samples confirmed the presence of this and other originally circulating “wild-type” variants, but documented only limited presence of the Beta variant and no other variants of concern [1–4]. This allowed a comparative assessment for the Alpha variant versus wild-type variants during this specific timeframe (Supplementary Text 3). From March 8 through May 31, 2021, the Beta variant wave expanded rapidly and viral genome sequencing [1–4] and multiplex quantitative reverse-transcription PCR (RT-qPCR) variant screening [1–5] indicated dominance of the Beta and Alpha variants, with limited presence of other variants [1–5]. This enabled comparisons between the Beta versus Alpha variants during this specific timeframe (Supplementary Text 3). The Delta [9] (B.1.617.2) variant has been introduced only recently in Qatar, and it remains at low incidence as of July 11, 2021 [3–5]. Further details on the classification of infections by variant type are found in Supplementary Text 3.

Descriptive statistics (frequency distributions and measures of central tendency) were used to characterize the study samples. Two-sided p-value of <0.05 was considered statistically significant. The odds ratio and its associated 95% confidence interval (CI) were calculated using the exact method. Confidence intervals were not adjusted for multiplicity. Interactions were not investigated. Two sensitivity analyses were conducted by first adjusting for age, and second by adjusting for age and sex, in logistic regression analyses. Statistical analyses were conducted in STATA/SE version 17.0.

The study was approved by the Hamad Medical Corporation and Weill Cornell Medicine-Qatar Institutional Review Boards with waiver of informed consent. Reporting of the study followed the STROBE guidelines (Supplementary Table 1).

## Results

Demographic characteristics of the samples for each disease outcome in assessing the severity, criticality, and fatality of the Alpha variant compared to the wild-type variants are presented in Supplementary Table 2. Compared to wild-type variants, the odds of progressing to severe disease were 1.48-fold (95% CI: 1.18-1.84) higher for the Alpha variant (Table 1). The odds of progressing to critical disease were 1.58-fold (95% CI: 0.79-3.10) higher, but did not reach statistical significance. There were too few COVID-19 deaths to assess the fatality of the Alpha variant.

**Table 1.** **Infection severity, criticality, and fatality of the Alpha and Beta variants in the population of Qatar**.

| Groups | Infection severity<sup>‡</sup> | Assessment of severity, criticality, and fatality of the Alpha variant compared to the wild-type variants circulating between January 18-February 15, 2021<sup>*</sup> |  |  | Assessment of severity, criticality, and fatality of the Beta variant compared to the Alpha variant between March 8-May 31, 2021<sup>†</sup> |  |  |
| --- | --- | --- | --- | --- | --- | --- | --- |
|  |  | Exposed | Not exposed | Odds ratio (95% CI) | Exposed | Not exposed | Odds ratio (95% CI) |
|  |  | Alpha variant | Wild-type variants |  | Beta variant | Alpha variant |  |
| Cases<sup>**</sup> | Severe disease | 188 | 279 | 1.48 (1.18-1.84) | 2,036 | 483 | 1.24 (1.11-1.39) |
| Controls<sup>**</sup> | Asymptomatic or mild infection | 431 | 944 |  | 5,806 | 1,707 |  |
| Cases<sup>**</sup> | Critical disease | 21 | 37 | 1.58 (0.79-3.10) | 382 | 81 | 1.49 (1.13-1.97) |
| Controls<sup>**</sup> | Asymptomatic or mild infection | 49 | 125 |  | 1,056 | 333 |  |
| Cases<sup>**</sup> | Severe or critical disease | 209 | 316 | 1.45 (1.18-1.79) | 2,418 | 564 | 1.28 (1.15-1.42) |
| Controls<sup>**</sup> | Asymptomatic or mild infection | 480 | 1,054 |  | 6,764 | 2,019 |  |
| Cases<sup>**</sup> | COVID-19 death | 2 | 9 | 0.83 (0.07-5.58) | 142 | 37 | 1.57 (1.03-2.43) |
| Controls<sup>**</sup> | Asymptomatic or mild infection | 7 | 26 |  | 381 | 156 |  |

Abbreviations: PCR, polymerase chain reaction.
<sup>\*</sup>From January 18-February 15, 2021, the Alpha variant and other wild-type variants dominated incidence with limited presence of the Beta variant[1-4].
<sup>†</sup>From March 8-May 31, 2021, the Beta and Alpha variants dominated incidence with limited presence for other variants[1-5].
<sup>‡</sup>Severe disease, critical disease, and COVID-19 death were defined based on the World Health Organization criteria for classifying SARS-CoV-2 infection severity[6] and COVID-19-related death[7].
<sup>\*\*</sup>Cases and controls were matched on a ratio of one-to-three by 10-year age group, sex, and bi-weekly interval of the PCR diagnosis date.

Demographic characteristics of the samples for each disease outcome in assessing the severity, criticality, and fatality of the Beta variant compared to the Alpha variant are presented in Supplementary Table 3. Compared to the Alpha variant, the odds of progressing to severe disease were 1.24-fold (95% CI: 1.11-1.39) higher for the Beta variant (Table 1). The odds of progressing to critical disease were 1.49-fold (95% CI: 1.13-1.97) higher. The odds of COVID-19 death were 1.57-fold (95% CI: 1.03-2.43) higher. Sensitivity analyses confirmed the above results (Supplementary Table 4).

## Discussion

The Alpha variant presented a 48% higher risk of severe disease than wild-type variants in the population of Qatar, affirming its greater gravity [10, 11] (odds ratio approximates risk ratio for rate outcomes). Infection with the Beta variant was associated with even greater risks of severe and critical disease and COVID-19 death, affirming earlier observational analyses suggesting its high gravity [11, 12]. Compared to the Alpha variant, infections with the Beta variant posed a 24% higher risk of severe disease, 49% higher risk of critical disease, and 57% higher risk of COVID-19 death.

These results explain the changing pattern of hospitalizations and deaths seen during the Beta wave compared to the Alpha wave (Figure 1). Acute-care admissions doubled during the Beta wave, but ICU admissions and deaths quadrupled, with the disproportionally greater effect of this variant on critical disease and COVID-19 death.

**Figure 1.**
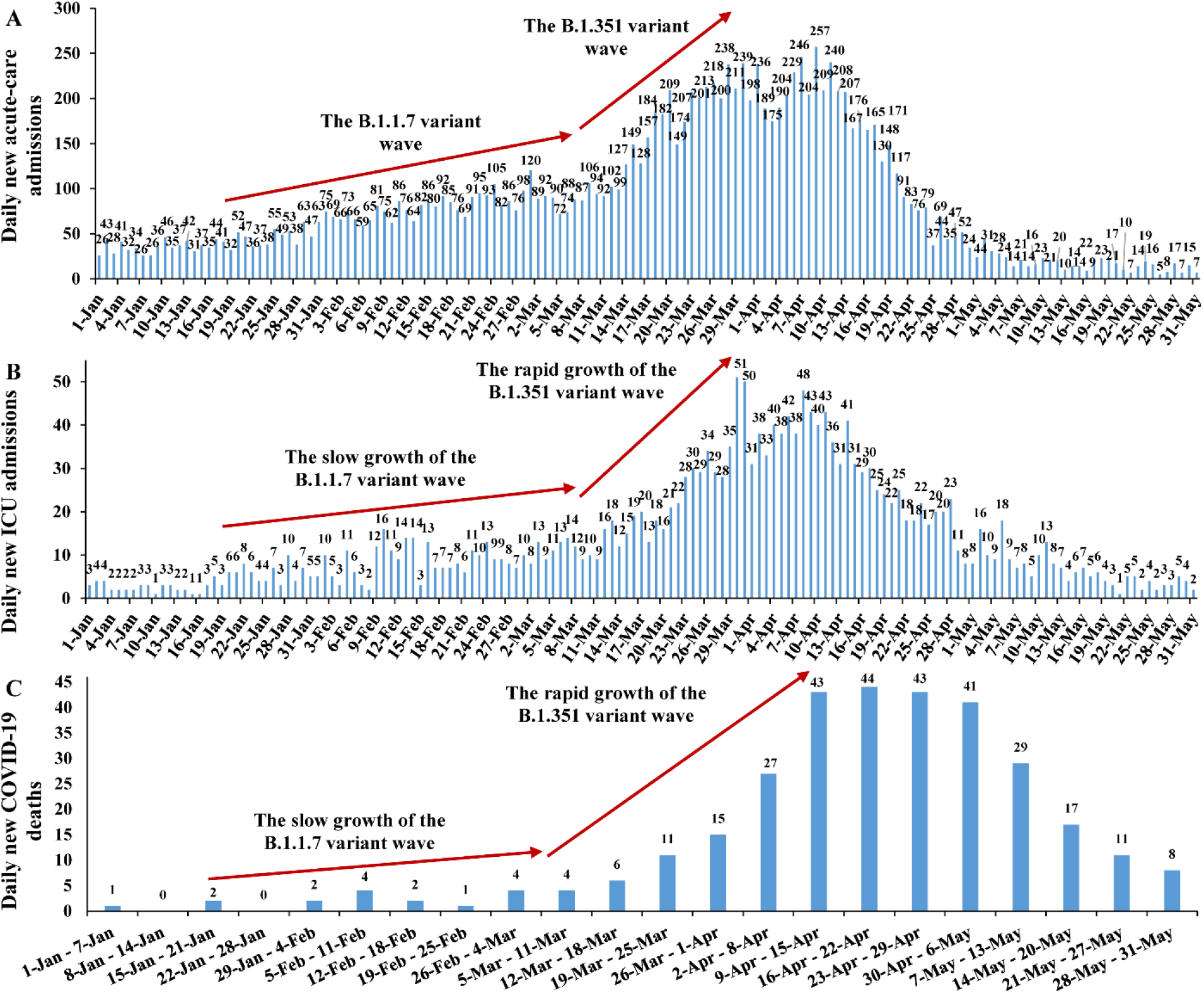
**Number of A) daily new COVID-19 acute-care hospital admissions, B) daily new COVID-19 ICU hospital admissions, and C) COVID-19 deaths in Qatar**.

Limitations include the smaller sample sizes of critical disease and COVID-19 deaths in the Alpha variant analysis (Supplementary Table 2) compared to the Beta variant analysis (Supplementary Table 3), as COVID-19 criticality and fatality have been low in Qatar’s predominantly young and working-age population [8, 13], leading to statistically non-significant results and wider 95% confidence intervals. Data on co-morbid conditions were not available to study investigators; hence, they could not be factored explicitly in our analysis. Nevertheless, matching and adjusting for age in analysis may have served as a proxy, given that co-morbidities are associated with old age. Furthermore, with the young population structure [8], we anticipate that only a small proportion of the study population may have had serious co-morbid conditions.

However, our findings may not be entirely generalizable to other settings, where elderly people constitute a sizable proportion of the population. Imperfect assay sensitivity and specificity of PCR testing may have affected infection ascertainment. However, all PCR testing was performed with extensively used, investigated, and validated commercial platforms having essentially 100% sensitivity and specificity (Supplementary Text 1). Unlike blinded, randomized clinical trials, the investigated observational cohorts were neither blinded nor randomized.

In conclusion, the Alpha variant is associated with 48% higher risk of severe disease than wild-type variants. In turn, the Beta variant is associated with 24% higher risk of severe disease than the Alpha variant, and strikingly, even higher risk of critical disease (49%) and COVID-19 death (57%). These findings highlight risks to healthcare systems, particularly intensive care facilities and resources, in the event of a globally increased circulation of the Beta variant.

## Data Availability

The dataset of this study is a property of the Qatar Ministry of Public Health that was provided to the researchers through a restricted-access agreement that prevents sharing the dataset with a third party or publicly. Future access to this dataset can be considered through a direct application for data access to Her Excellency the Minister of Public Health (https://www.moph.gov.qa/english/Pages/default.aspx). Aggregate data are available within the manuscript and its Supplementary information.

## Acknowledgements

We acknowledge the many dedicated individuals at Hamad Medical Corporation, the Ministry of Public Health, the Primary Health Care Corporation, and the Qatar Biobank for their diligent efforts and contributions to make this study possible.

## Author contributions

LJA conceived and co-designed the study, led the statistical analyses, and co-wrote the first draft of the article. HC co-designed the study, performed the statistical analyses, and co-wrote the first draft of the article. All authors contributed to data collection and acquisition, database development, discussion and interpretation of the results, and to the writing of the manuscript.

All authors have read and approved the final manuscript.

## Supplementary Material

### Supplementary Text 1. Laboratory methods

Nasopharyngeal and/or oropharyngeal swabs (Huachenyang Technology, China) were collected for PCR testing and placed in Universal Transport Medium (UTM). Aliquots of UTM were: extracted on a QIAsymphony platform (QIAGEN, USA) and tested with real-time reverse-transcription PCR (RT-qPCR) using TaqPath™ COVID-19 Combo Kits (100% sensitivity and specificity [1]; Thermo Fisher Scientific, USA) on an ABI 7500 FAST (ThermoFisher, USA); extracted using a custom protocol [2] on a Hamilton Microlab STAR (Hamilton, USA) and tested using AccuPower SARS-CoV-2 Real-Time RT-PCR Kits (100% sensitivity and specificity [3]; Bioneer, Korea) on an ABI 7500 FAST; or loaded directly into a Roche cobas® 6800 system and assayed with a cobas® SARS-CoV-2 Test (95% sensitivity, 100% specificity [4]; Roche, Switzerland). The first assay targets the viral S, N, and ORF1ab regions. The second targets the viral RdRp and E-gene regions, and the third targets the ORF1ab and E-gene regions.

All PCR tests were conducted at the Hamad Medical Corporation Central Laboratory or Sidra Medicine Laboratory, following standardized protocols.

### Supplementary Text 2. COVID-19 severity, criticality, and fatality classification

Severe COVID-19 disease was defined per WHO classification as a SARS-CoV-2 infected person with “oxygen saturation of <90% on room air, and/or respiratory rate of >30 breaths/minute in adults and children >5 years old (or ≥60 breaths/minute in children <2 months old or ≥50 breaths/minute in children 2–11 months old or ≥40 breaths/minute in children 1–5 years old), and/or signs of severe respiratory distress (accessory muscle use and inability to complete full sentences, and, in children, very severe chest wall indrawing, grunting, central cyanosis, or presence of any other general danger signs)” [5]. Detailed WHO criteria for classifying SARS-CoV-2 infection severity can be found in the WHO technical report [5].

Critical COVID-19 disease was defined per WHO classification as a SARS-CoV-2 infected person with “acute respiratory distress syndrome, sepsis, septic shock, or other conditions that would normally require the provision of life sustaining therapies such as mechanical ventilation (invasive or non-invasive) or vasopressor therapy” [5]. Detailed WHO criteria for classifying SARS-CoV-2 infection criticality can be found in the WHO technical report [5].

COVID-19 death was defined per WHO classification as “a death resulting from a clinically compatible illness, in a probable or confirmed COVID-19 case, unless there is a clear alternative cause of death that cannot be related to COVID-19 disease (e.g. trauma). There should be no period of complete recovery from COVID-19 between illness and death. A death due to COVID-19 may not be attributed to another disease (e.g. cancer) and should be counted independently of preexisting conditions that are suspected of triggering a severe course of COVID-19”. Detailed WHO criteria for classifying COVID-19 death can be found in the WHO technical report [6].

### Supplementary Text 3. Classification of infections by variant type

Classification of infections by variant type was informed by weekly rounds of viral genome sequencing and multiplex, real-time reverse-transcription PCR (RT-qPCR) variant screening [7] of randomly collected clinical samples [8–12], as well as by results of deep sequencing of wastewater samples [10]. Based on existing evidence [13–15] and confirmation with viral genome sequencing [16], an Alpha case was defined as an S-gene “target failure” using the TaqPath COVID-19 Combo Kits (Thermo Fisher Scientific, USA [1]; >85% of PCR testing in Qatar) applying the criterion of a PCR cycle threshold (Ct) value ≤30 for both the N and ORF1ab genes, and a negative outcome for the S gene [15].

From January 18 until February 15, 2021, the weekly rounds of viral genome sequencing [8–11] identified presence of the Alpha variant and other originally circulating “wild-type” variants, but documented only limited presence of the Beta variant and no presence of other variants of concern [8–11]. Meanwhile, with essentially only Beta and Alpha cases identified from March 8, 2021 until the end of study (May 31, 2021) in the viral genome sequencing and multiplex RT-qPCR variant screening [8–12], a Beta case was proxied as the complement of Alpha criteria, that is, any infection with a Ct value ≤30 for the N, ORF1ab, and S genes. The Delta [17] (B.1.617.2) variant has been introduced only recently in Qatar, and it remains at low incidence as of July 11, 2021 [10–12]. There is no evidence that any other variant of concern is or has been responsible for appreciable community transmission in Qatar [10, 11].

**Supplementary Table 1.** STROBE checklist for case-control studies.

|  | Item No | Recommendation | Main text page # |
| --- | --- | --- | --- |
| Title and abstract | 1 | (a) Indicate the study’s design with a commonly used term in the title or the abstract | 2 |
|  |  | (b) Provide in the abstract an informative and balanced summary of what was done and what was found | 2 |
| **Introduction** |  |  |  |
| Background/rationale | 2 | Explain the scientific background and rationale for the investigation being reported | 3 |
| Objectives | 3 | State specific objectives, including any prespecified hypotheses | 3 |
| **Methods** |  |  |  |
| Study design | 4 | Present key elements of study design early in the paper | 3-4 |
| Setting | 5 | Describe the setting, locations, and relevant dates, including periods of recruitment, exposure, follow-up, and data collection | 3-4 |
| Participants | 6 | (a) Give the eligibility criteria, and the sources and methods of case ascertainment and control selection. Give the rationale for the choice of cases and controls | 3-4 & Supp. Text 2 |
|  |  | (b) For matched studies, give matching criteria and the number of controls per case | 4 |
| Variables | 7 | Clearly define all outcomes, exposures, predictors, potential confounders, and effect modifiers. Give diagnostic criteria, if applicable | 3-5 & Supp. Texts 1-3 |
| Data sources/measurement | 8* | For each variable of interest, give sources of data and details of methods of assessment (measurement). Describe comparability of assessment methods if there is more than one group | 3, 5 & Supp. Texts 1-3 |
| Bias | 9 | Describe any efforts to address potential sources of bias | 4-5 |
| Study size | 10 | Explain how the study size was arrived at | 3-4 |
| Quantitative variables | 11 | Explain how quantitative variables were handled in the analyses. If applicable, describe which groupings were chosen and why | Table 1 & Supp. Tables 2-4 & Supp. Text 3 |
| Statistical methods | 12 | (a) Describe all statistical methods, including those used to control for confounding | 4-5 |
|  |  | (b) Describe any methods used to examine subgroups and interactions | NA |
|  |  | (c) Explain how missing data were addressed | NA, see p. 3 |
|  |  | (d) If applicable, explain how matching of cases and controls was addressed | 4 |
|  |  | (e) Describe any sensitivity analyses | 5 |
| **Results** |  |  |  |
| Participants | 13* | (a) Report numbers of individuals at each stage of study—eg numbers potentially eligible, examined for eligibility, confirmed eligible, included in the study, completing follow-up, and analysed | 3-5, Table 1 & Supp. Tables 2-3 |
|  |  | (b) Give reasons for non-participation at each stage | NA, see p. 3 |
|  |  | (c) Consider use of a flow diagram | NA |
| Descriptive data | 14* | (a) Give characteristics of study participants (eg demographic, clinical, social) and information on exposures and potential confounders | 5, Supp. Tables 2-3 |
|  |  | (b) Indicate number of participants with missing data for each variable of interest | NA, see p. 3 |
| Outcome data | 15* | Report numbers in each exposure category, or summary measures of exposure | 5-6, Table 1 & Supp. Table 4 |
| Main results | 16 | (a) Give unadjusted estimates and, if applicable, confounder-adjusted estimates and their precision (eg, 95% confidence interval). Make clear which confounders were adjusted for and why they were included | 5-6 & Table 1 |
|  |  | (b) Report category boundaries when continuous variables were categorized | Table 1 & Supp. Tables 2-4 |
|  |  | (c) If relevant, consider translating estimates of relative risk into absolute risk for a meaningful time period | NA |
| Other analyses | 17 | Report other analyses done—eg analyses of subgroups and interactions, and sensitivity analyses | 6 & Supp. Table 4 |
| **Discussion** |  |  |  |
| Key results | 18 | Summarise key results with reference to study objectives | 6 |
| Limitations | 19 | Discuss limitations of the study, taking into account sources of potential bias or imprecision. Discuss both direction and magnitude of any potential bias | 6-7 |
| Interpretation | 20 | Give a cautious overall interpretation of results considering objectives, limitations, multiplicity of analyses, results from similar studies, and other relevant evidence | 7 |
| Generalisability | 21 | Discuss the generalisability (external validity) of the study results | 7 |
| **Other information** |  |  |  |
| Funding | 22 | Give the source of funding and the role of the funders for the present study and, if applicable, for the original study on which the present article is based | 8 |

NA, not applicable; Supp, supplementary

**Supplementary Table 2.**
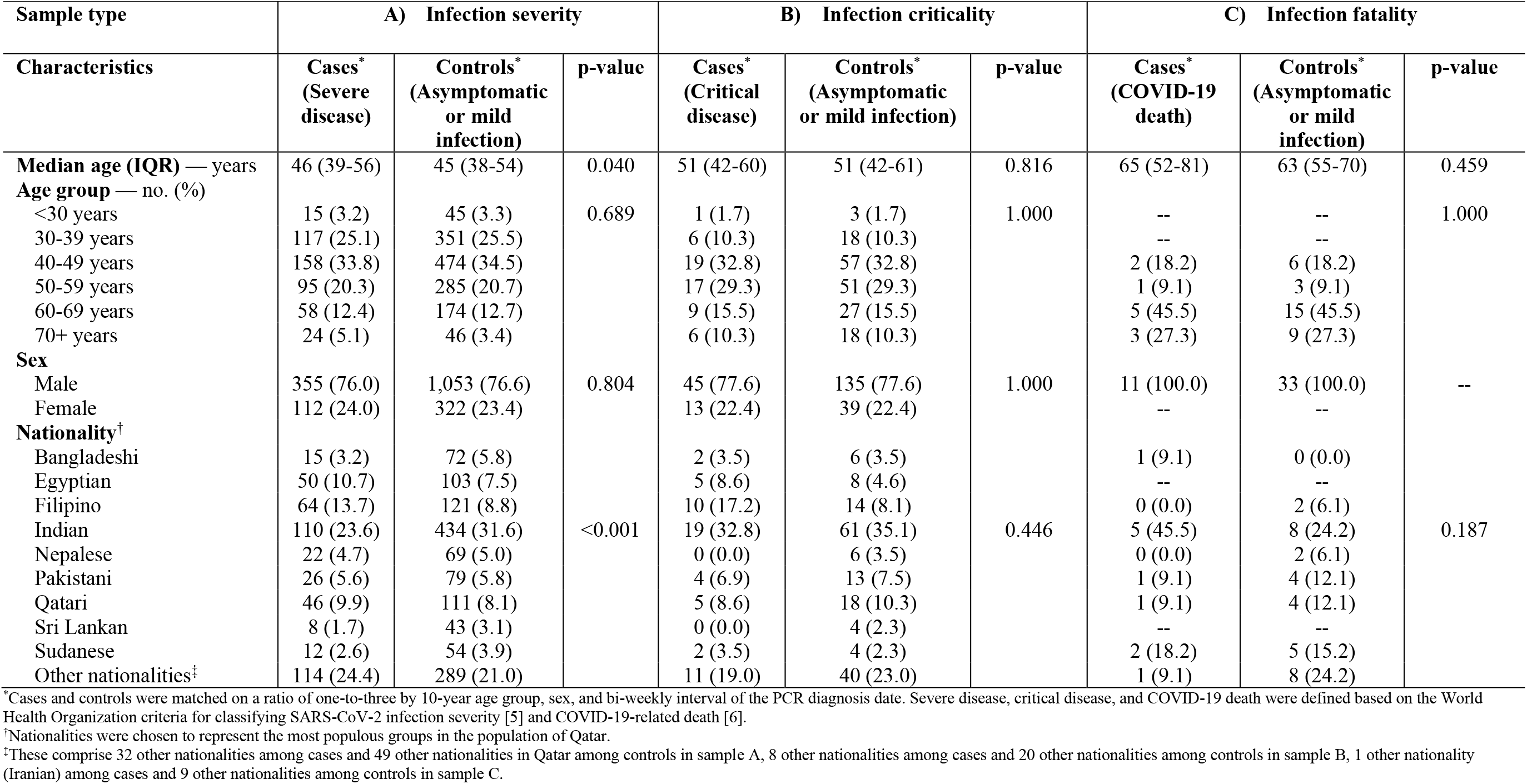
Assessment of the severity, criticality, and fatality of the Alpha variant compared to wild-type variants. Demographic characteristics of A) patients experiencing COVID-19 severe disease and controls (asymptomatic or mild SARS-CoV-2 infection), B) patients experiencing COVID-19 critical disease and controls (asymptomatic or mild SARS-CoV-2 infection), and C) cases resulting in COVID-19 death and controls (non-fatal infection).

**Supplementary Table 3.**
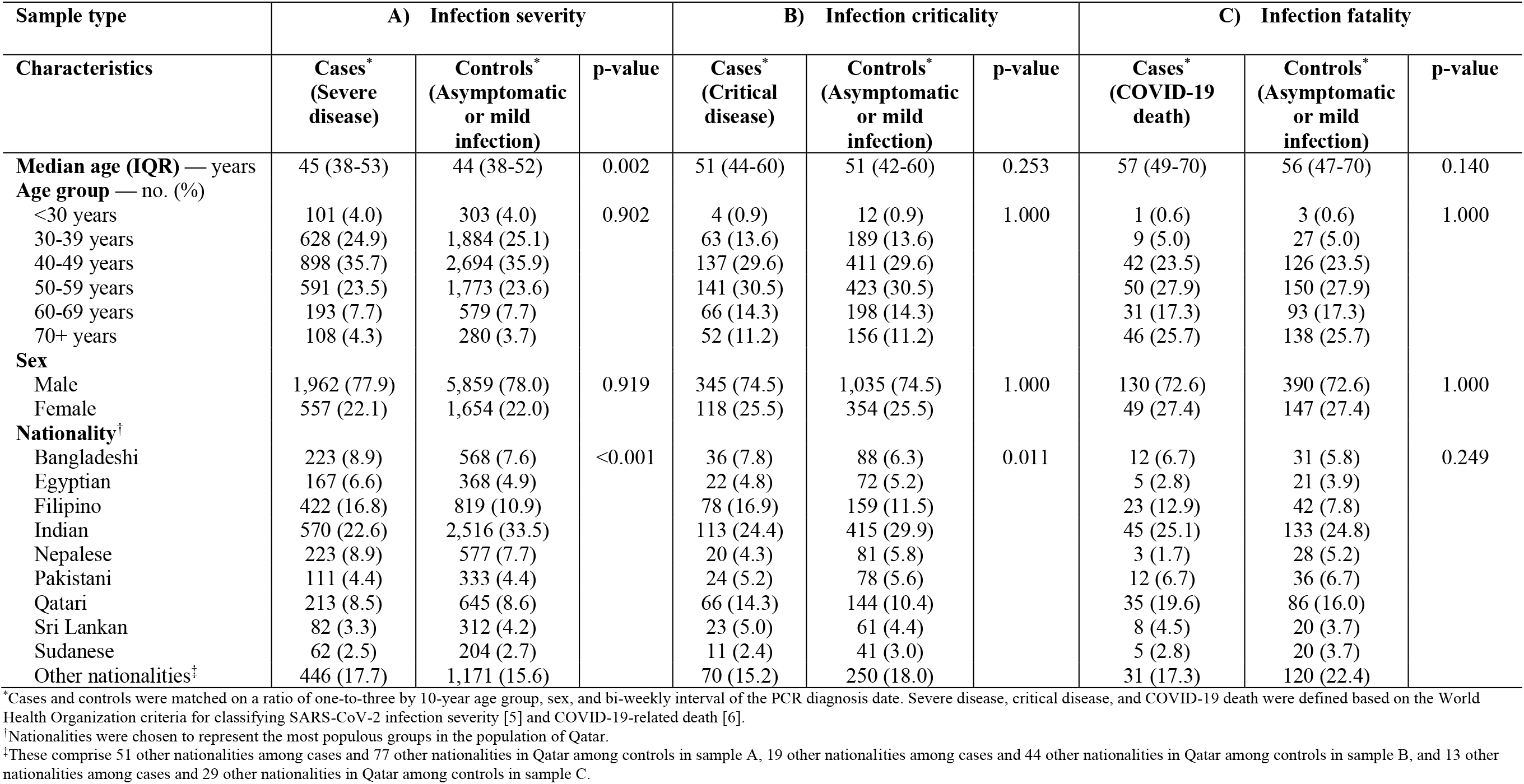
Assessment of the severity, criticality, and fatality of the Beta variant compared to the Alpha variant. Demographic characteristics of A) patients experiencing COVID-19 severe disease and controls (asymptomatic or mild SARS-CoV-2 infection), B) patients experiencing COVID-19 critical disease and controls (asymptomatic or mild SARS-CoV-2 infection), and C) cases resulting in COVID-19 death and controls (non-fatal infection).

**Supplementary Table 4.**
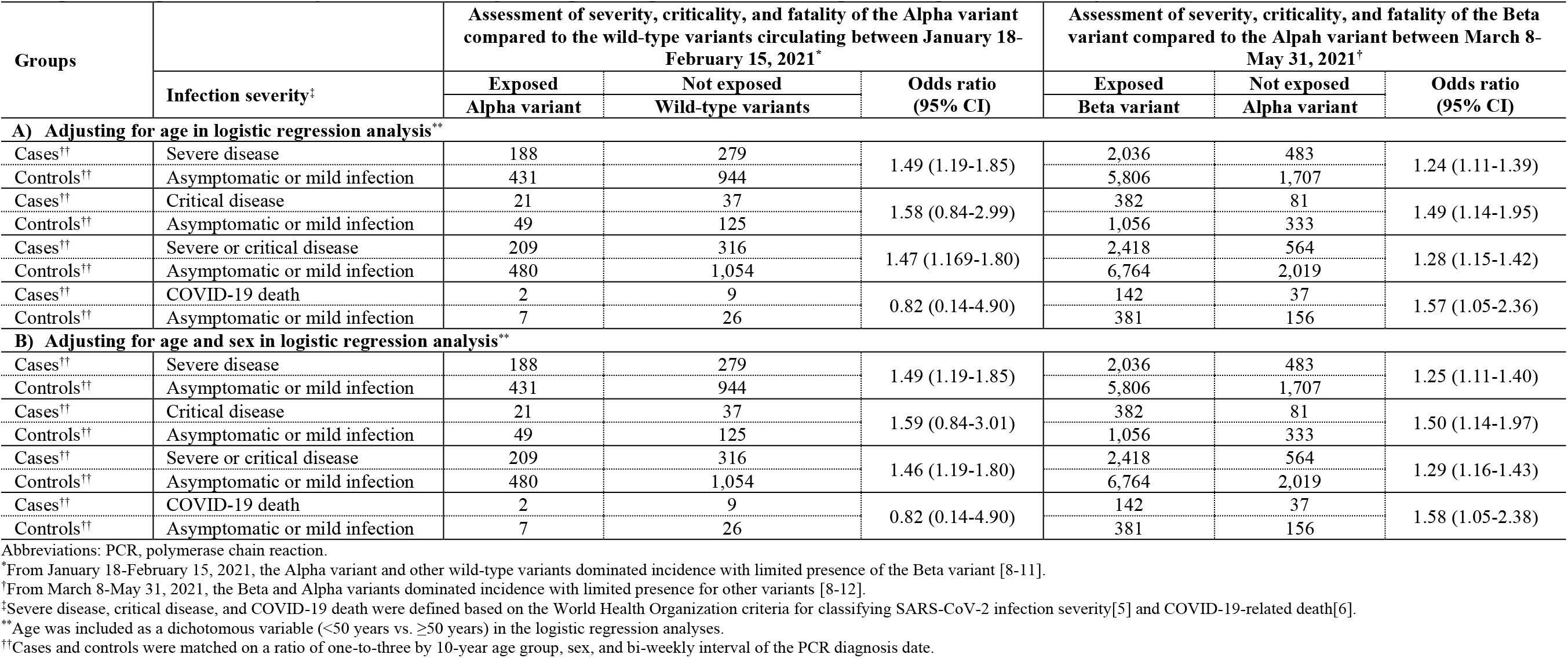
Sensitivity analyses for COVID-19 severity, criticality, and fatality in Qatar after A) adjusting for age in logistic regression analysis, and B) adjusting for age and sex in logistic regression analysis.

